# Yearly trends in cause-specific age-adjusted mortality from 1999-2020 in Black and White Non-Hispanic Americans

**DOI:** 10.1101/2024.10.01.24314642

**Authors:** Adith S. Arun, Harlan M. Krumholz

## Abstract

From 1999 to 2020, disparities in all-cause age-adjusted mortality rates between Black and White Non-Hispanic Americans decreased then stagnated and increased. Consistent improvement in racial disparities were made for cancer, HIV, and sepsis regardless of all-cause trends. In contrast, conditions like heart disease and external causes of death mirrored the overall trends.

---

Non-Hispanic Black Americans experienced higher all-cause age-adjusted mortality rates (AAMR) than Non-Hispanic White Americans from 1999-2020^1^. The excess all-cause age-adjusted mortality rate (eAAMR), defined as the Black AAMR minus the White AAMR, revealed a period of declining excess mortality rates followed by a period of stagnation and increase^1,2^. For males, eAAMRs declined from 1999-2011 and did not decrease from 2012-2020^1,2^. For females, eAAMRs declined from 1999-2015 and did not decrease from 2016-2020^1,2^.

Consistent eAAMR declines in cancer, HIV, and sepsis occurred during both periods of decline and no decline for males (Fig. 1). On the other hand, heart disease, diabetes, accidents, assaults, and suicide eAAMRs were concordant with the all-cause eAAMR trends (Fig. 1). They did not increase during the period of decline and did not decrease during the period of no decline. For females, consistent eAAMR declines irrespective of the all-cause period were similarly observed in cancer, HIV, and sepsis (Fig. 2). Relative to males, the eAAMR increases in specific causes were less pronounced for females during the period of no decline in all-cause eAAMRs (Fig. 2). Increases in eAAMRs from 2016-2020 were still observed in most causes including heart disease, cerebrovascular diseases, diabetes, accidents, suicides, and assaults (Fig. 2). For both males and females, eAAMRs increased markedly for most causes in 2020, the start of the pandemic, compared with 2019. The pandemic-related exacerbation of racial disparities has been characterized in other work^3^. The eAAMRs were largest for heart disease, cancer, HIV and assault in males whereas for females, eAAMRs were largest for heart disease, cancer, and diabetes. eAAMRs were negative—higher AAMRs for White relative to Black Americans—for a few conditions including Parkinson’s disease and chronic liver disease (Fig. 1, 2).

**Figure 1.**
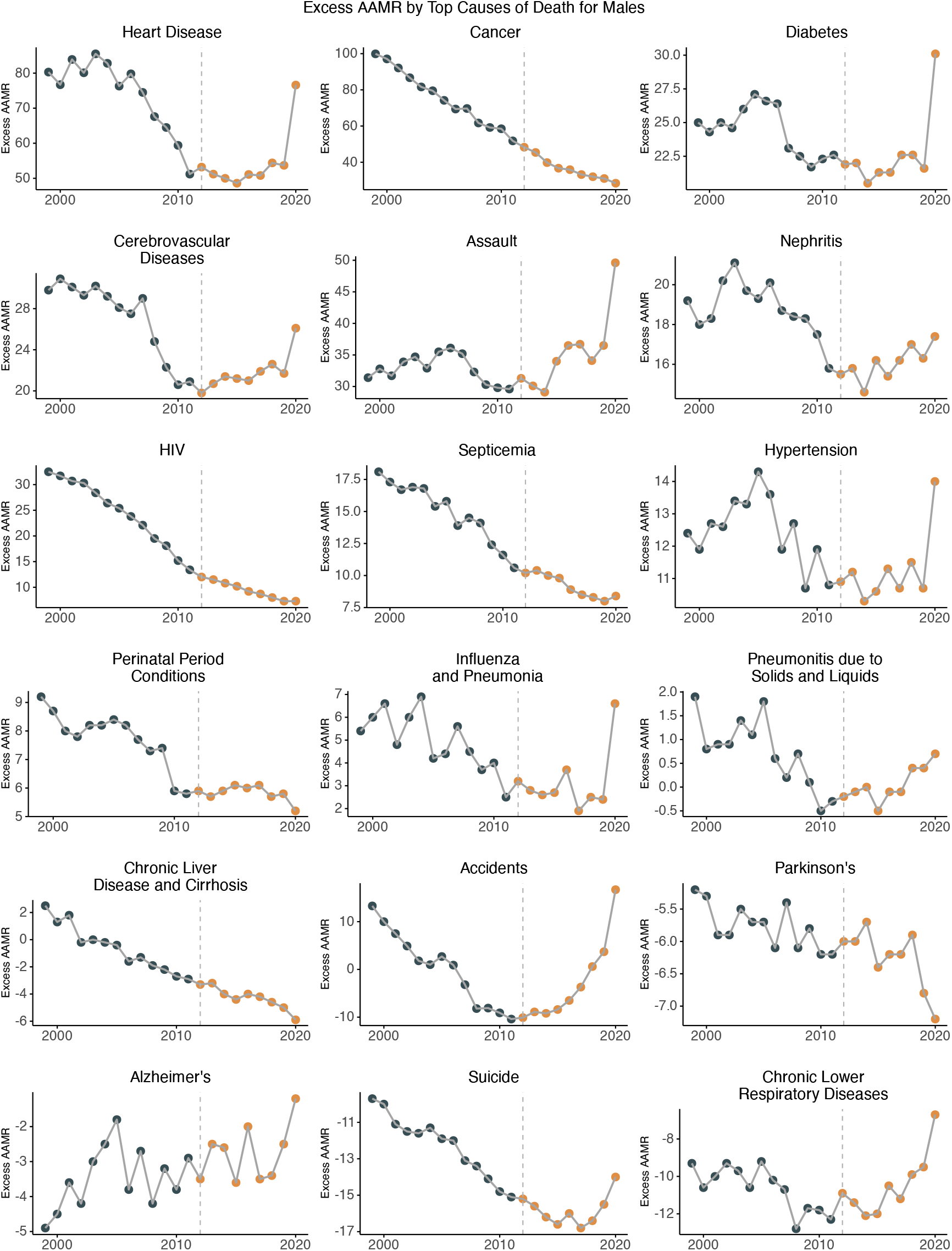
Cause-specific excess age-adjusted mortality rates (eAAMRs) for males from 1999-2020. The navy dots indicate the period of decline in all-cause eAAMRs for males (1999-2011); orange dots indicate the period of no decline in all-cause eAAMRs (2012-2020).

**Figure 2.**
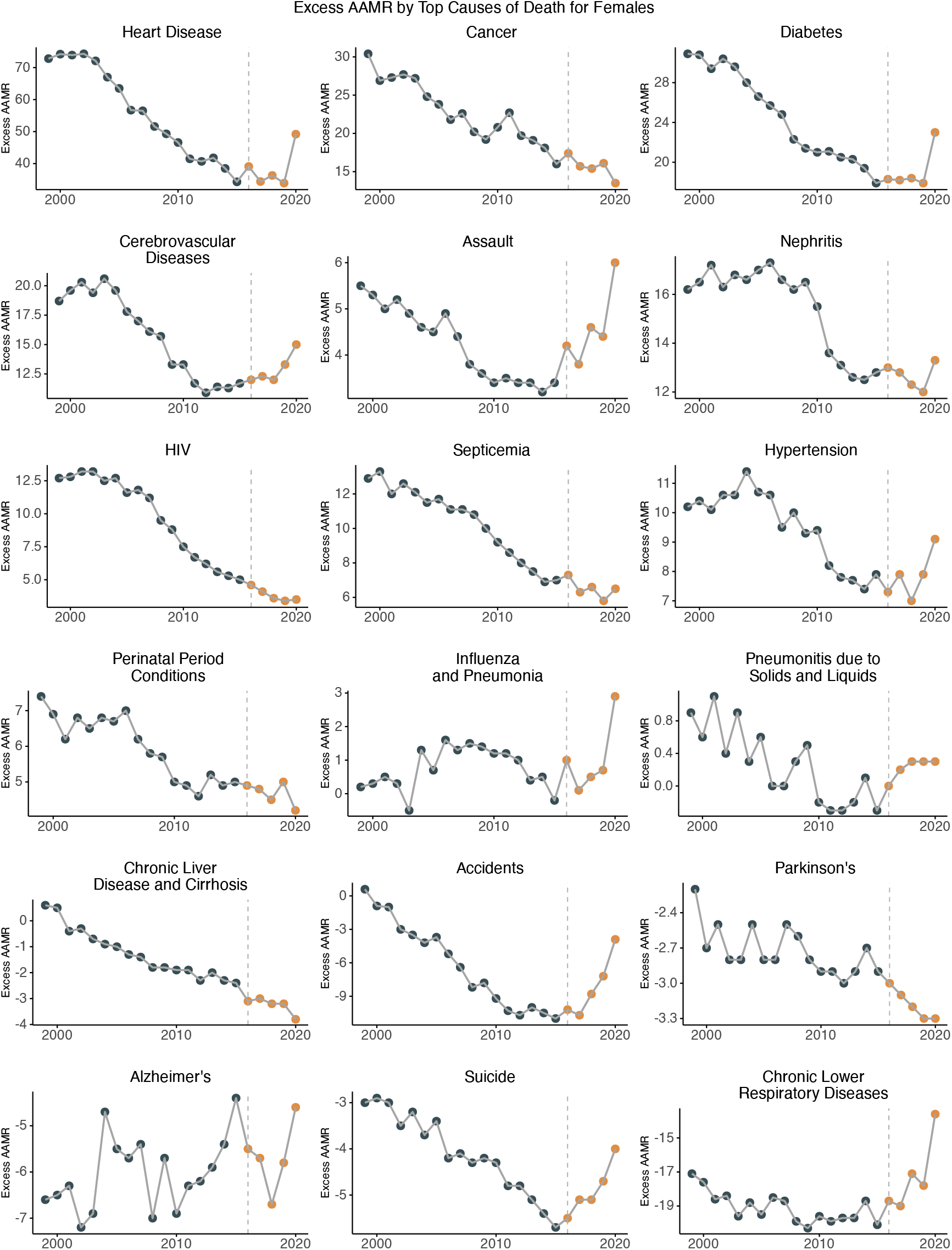
Cause-specific excess age-adjusted mortality rates (eAAMRs) for females from 1999-2020. The navy dots indicate the period of decline in all-cause eAAMRs for females (1999-2015); orange dots indicate the period of no decline in all-cause eAAMRs (2016-2020).

Visualizing cause-specific yearly eAAMRs in the context of all-cause changes allows us to quickly understand specific diseases in which progress towards lessening racial disparities may or may not be occurring. Since 2012 and 2016 for males and females respectively, Black Americans have experienced markedly worse outcomes in external causes of death (accidents, assaults, and suicides). Possible reasons for this change include inflation, unstable employment, and food insecurity but further work is needed to address this crisis both at the level of policy and research^4^. Further, these trends indicate that there are pockets of continued improvement in racial disparities, specifically cancer-related outcomes, regardless of the all-cause context. This suggests that we may be able to identify factors in cancer-related care that could be applied to heart disease and other causes to make progress on racial disparities.

Age-adjusted mortality rates were collected from US Centers for Disease Control and Prevention (CDC) Wide-Ranging Online Data for Epidemiologic Research (WONDER) death certificates. Data and code to replicate these analyses are publicly available at https://github.com/aditharun/us-mort-disp-analysis.

## Data Availability

Data is publicly available from wonder.cdc.gov and code to analyze data is publicly available at https://github.com/aditharun/us-mort-disp-analysis

## Additional Contributions

We thank Drs. César Caraballo, Mitsuaki Sawano, Yuan Lu, Rohan Khera, Clyde W. Yancy, Frederick Warner, and Jeph Herrin for their scientific perspective and comments on this work.

